# Epilepsy in children with perinatal arterial ischemic stroke

**DOI:** 10.1101/2021.09.29.21263933

**Authors:** Fabienne Kühne, Alexander Jungbluth, Joanna Schneider, Christoph Bührer, Christine Prager, Angela M. Kaindl

## Abstract

**Purpose:** Perinatal ischemic stroke (PIS) is a frequent cause for perinatal brain structure defects resulting in epilepsy, cerebral palsy and disability. Since the severity of symptoms is variable, the aim of this study was to evaluate the outcome of children with PIS and seizures/epilepsy to aid parental counseling and therapy decisions.

**Material:** We studied retrospectively patients with arterial PIS and structural epilepsy or seizures in the newborn treated at a single center in 2000-2019. Specifically, signs and symptoms of cerebral palsy (CP), developmental and motor delay, epilepsy and thrombophilia were assessed.

**Results:** From the identified 69 individuals with arterial PIS, we only included the 50 patients (64% male) who had structural epilepsy at the time of investigation or previously in their medical history.The mean age of the included patients was 7.1 years (range 0.08-22) at last consultation. Infarct localisation was predominantly unilateral (86%), left sided (58%) and affecting the middle cerebral artery (94%). Genetic thrombophilia was identified in 52% of the patients examined with genetic testing. More than half of the individuals had CP (52%), and 38.5% had a cognitive outcome below average. First seizures occurred in the neonatal period in 58% of patients and developed into drug-refractory epilepsy in 24.1%. Children with late-onset of epilepsy were twice as likely to develop drug-refractory epilepsy (52.4%).

**Discussion:** Our study shows that patients with PIS and seizures as common sequela often also develop CP. Children with later onset of epilepsy have a worse outcome. Patients with seizure onset in the neonatal period and reccuring seizures have a good response to treatment. Therefore, early diagnosis, follow-up examination and adequate therapy are important. Most children need intensive physiotherapy and speech therapy; however, participation in life is usually age-appropriate.

## INTRODUCTION

Perinatal ischemic stroke (PIS) occurs with an incidence of 1:2300-4000 live-births and is an important cause of live-long neurologic morbidity including cerebral palsy (CP), epilepsy and intellectual disability^1–4^. PIS is defined as a cerebrovascular event that occurs during pregnancy and up to 28 days postnatally^5^. PIS can be differentiated by the timing of presentation (in neonatal period versus later) and the vascular lesion (arterial versus venous)^6^. During pregnancy PIS can be an incidental finding in routine ultrasound examinations^2^. Prenatal counselling is difficult due to limited knowledge on the developmental impact of a PIS. Postnatally, some affected children rapidly develop neurologic symptoms such as neonatal seizures, while others do not have any abnormalities within the neonatal period and may be diagnosed retrospectively when hemiparesis, seizures, developmental delay or other neurological symptoms are investigated by neuroimaging later in life^4,7–12^. Epilepsy, an important sequela of PIS, occurs with an incidence of 16-39% in individuals with PIS and holds a wide range in response to treatment^3,7,9,13–15^. Neonatal seizures often show a good response to antiepileptic treatment, and a late-onset epilepsy as a result of a PIS is often accompanied by a developmental delay^5,15–20^.

Thus, parental counseling remains difficult even postnatally given the limited knowledge on the impact of PIS on neurodevelopment. The aim of this study was to evaluate the neurologic outcome of children with PIS and seizures/epilepsy to aid parental counseling and therapeutic decisions.

## SUBJECTS AND METHODS

### Subjects

We studied retrospectively patients with PIS who were examined in a standardized manner at the Center for Chronically Sick Children of Charité University Medicine Berlin, Germany, between January 2000 and March 2019. Patients were identified through a systematic digital search using the search terms “perinatal stroke”, “perinatal arterial stroke”, “perinatal cerebral infarction”, “neonatal stroke”; and “neonatal cerebral infarction” in the electronic patient database SAP-CSP (SAP SE, Walldorf, Germany). The information was extracted from medical paper-based and electronic patient files using an electronic data-collection template that had been devised specifically for this cross-sectional study: demographic data (age, gender, ethnicity), medical history including gestational week (GW) at birth, family history, stroke location, neurological findings, radiological examinations (cranial magnetic resonance imaging (MRI) and cranial ultrasounds), genetic test results for thrombophilia, neurophysiological testing including EEG, seizure semiology, anticonvulsant drugs, effectiveness of drugs, developmental data, socio-demographic and psycho-social aspects. Cranial MRIs of patients with PIS were reanalyzed by FK. The study was approved by the local ethics committee and data security commission (no. EA2/084/18).

From the identified 69 individuals with arterial PIS, we included only the 50 patients who had acute symptomatic seizures in the newborn or had structural epilepsy at the time of investigation or previously in their medical history. 19 patients with PIS where excluded as they did not develop seizures within a mean follow-up time of 5.8 years. Most of these patients where diagnosed having had a PIS through MRI scans initiated for postnatal complications (15/19), including postnatal resuscitation (3/19), or later apparent paresis (4/19).

### Thrombophilia screening

In the diagnostic workup of an individual with PIS tests for thrombophilia were performed routinely at our center to rule out such causes. Specifically, blood sampling was performed in order to determine the following factors: increased platelet count, shortening of the coagulation time (aPTT, PT), elevations of factor VIII (FVIII), factor XII (FXII), fibrinogene (Fg) and lipoprotein (a) (Lpa), decreased, protein C (PC), protein S (PS), antithrombin (ATIII) as well as genetic test for hetereo- and homocygotic factor V Leiden mutation (mutation in G1691A or F506Q), prothrombin gene mutation G20210A and PAI-1 polymorphisms. Moreover, consanguinity of the patient’s parents and a positive family history for thrombophilia was assessed.

### Assessment of developmental outcome

The development outcome of the patients was assessed based on medical records. This included data on cognition, speech and motor function development.

The cognitive outcome was defined as below average, average and above average, based on the total IQ score: normal cognitive development (‘average’) was defined by an IQ of 85-115, intellectual disability (‘below average’) by IQ scores below 85 or when testing was impossible due to severe intellectual disability and ‘above average’ by IQ scores above 115. Cognition was examined using conventional age-appropriate tests such as Bayley-Scales of Infant development II at 1 to 4 years of age Kaufman Assessment Battery for Children (K-ABC) 1^st^ edition for children aged 3 to 18 years and Wechsler Intelligence Scales for Children (WISC-IV) 4th edition for children aged 6 to 16 years. Patients with no cognitive testing available were categorized based on clinical assessment data.

Speech development was grouped as normal, moderately and severely delayed, in which mutism and an inability to speak multi-word sentences at the age of 6 years was classified as severe speech development delay. Patients were classified as moderately delayed when they obtained speech therapy or did not achieve two-word-sentence-structures.

Motor development was evaluated via the achievement of the motor milestones. Additionally, in patients with CP the Gross Motor Function Classification System (GMFCS) was used which includes information like the muscle tone, the requirement of physiotherapy or orthoses^21^.

### Statistical analysis

The cumulate data was imported into the statistics program SPSS (version 25) for analysis and evaluation. The Fisher exact test was performed in 2×2 contingency tables with few expected frequencies. Additionally Mann-Whitney U test was done. Test results with a p-value < 0.05 were considered statistically significant. Figures were generated with GraphPad Prism version 7.0 (GraphPad Software, La Jolla, CA, USA).

## RESULTS

The study cohort comprised, from an initial cohort of 69 individuals with arterial PIS, a total of 50 individuals (32 male, 18 female) with arterial PIS <u>and</u> structural epilepsy or acute symptomatic seizures in the newborn at a mean age of 7.1 years (range 0.08-22) at last consultation (**Table 1**). (**Figure A**). The majority of them was born at term (40/50, 80% term; 7/50, 14% preterm with median GW 37 (range 32-37); 3/50, 6% unknown GW) and via vaginal delivery (30/50, 60%) to mothers with a median age of 31.5 years (range 15-42). Perinatal complications such as asphyxia and bradycardia occurred in almost a third (30%, n=15), 10.5% (4/38) had a cord blood pH <7.10, and 4% (2/50) required resuscitation. The Apgar scores at five and ten minutes were <7 in 22.2% (10/45) and 20% (9/45), and <5 in 13.3% (6/45) and 11.1% (5/45), respectively. At birth, the occipito-frontal head circumference was low-normal with a median of 34 cm (range 27.5-37) corresponding to a mean SD of 1.93, mean age-corrected percentile for head circumference was 31.6 P. (range <1-97 P.) while medians of weight (3245g, range 1720-4400) and length (50 cm, range 40.5-56) were normal.

Results of extended thrombophilia analysis that is performed routinely in all children with stroke at our center were available for 41 subjects. Here, abnormalities were present in 43.9% (18/41) of cases, detailed in **Table 1**. Abnormal results of genetic thrombophilia tests in 52% (13/25) included 7 cases with factor V Leiden mutations, 2 cases with prothrombin mutations and 4 cases with PAI-1 polymorphisms. Familial thrombophilia was present in two cases, one with a prothrombin mutation, another with a factor V mutation. Two siblings were affected by a stroke, one of them with a factor V Leiden mutation. Two cases were children of consanguine healthy parents but did not carry a variant in any of the mentioned thrombophilia genes.

We further analyzed the radiologic features of patients with PIS (**Table 1**). The timing of neuroimaging depended on that of first symptoms: neonatal-onset seizures lead to neuroimaging within the first days of life in 29 cases. Primary neuroimaging in all of these patients was cranial ultrasound, followed by cranial MRI in 89.6% (26/29) and by cranial CT in one outborn case. MRI was not applicable in one case due to a pacemaker for third-degree AV block, and in another outborn case there was no MRI available postnatally. MRI scans were performed in all patients (n=21) who were asymptomatic as neonates but developed structural epilepsy later. The majority (86%, 43/50) of children had a unilateral PIS, affecting predominantly the left hemisphere (58% left, 28% right). Bilateral stroke had occurred in 7/50 cases (14%). In nearly all cases the stroke affected the middle cerebral artery (MCA; 94%, 47/50), followed by the posterior cerebral artery (PCA; 16%, 8/50), lacunar strokes in the basal ganglia (13%, 7/50) and strokes affecting the anterior cerebral artery (ACA, 3/50, 4.3%) (**Table 1**). In addition, 10 cases had signs of hemorrhage on MRI (petechial bleedings in 7 and parenchymal hematomas in 3 cases). Although 49 mothers (98%) had received regular pregnancy check-ups, only one case of PIS had been identified prenatally.

### Epilepsy and anticonvulsant therapy

The onset of seizures demonstrates a two-peaked course. More than half of the patients developed acute symptomatic seizures in the neonatal period 58% (n= 29) (**Figure A**). The mean age at the first acute symptomatic seizure in the newborn was 2 days with a range from the first to the twelfth day of life. Nearly all acute symptomatic seizures in the newborn (85.7%) were focal in onset with awareness. Only three patients showed generalized-onset seizures, and one had focal-onset seizures with loss of awareness. Twenty-one patients (42%) experienced their first seizure after the neonatal period at a mean age of 4 years (range 0.2 to 12 years). Onset was within the first year of life but after the neonatal period in eight cases (38.1%) and beyond the first year of life in 13 cases (61.9%). Of all non-neonatal seizures with known semiology, 46.2% (n=12) were focal-onset with awareness, 38.5% generalized-onset and 15.3% focal-onset without awareness. At least one status epilepticus occurred in eight patients (16%), thereof three patients (37.5%) with acute symptomatic seizures in the newborn and five (62.5%) with onset in later childhood. EEGs could be reviewed in 40 cases. 32 (80%) of those showed epileptic discharges (23 focal, 3 focal with secondary generalization, 3 multifocal, 4 generalized).

All patients with arterial PIS and seizures were treated with at least one anticonvulsant drug in their lifetime. Cases with acute symptomatic seizures in the newborn were almost all first treated with phenobarbital (89.7%, 26/29). Two patients had self-limiting seizures, and applied drug was unknown in one case. More than half of the cases with acute symptomatic seizures in the newborn (51.7%, 15/29) were seizure-free at the time of study and did not require any further anticonvulsant drug (mean age at the end of the study 5.5 years). However, about a fourth of the cases with acute symptomatic seizures in the newborn (24.1%, 7/29) developed drug-refractory epilepsy, i.e., therapy with two or more anticonvulsive drugs had failed to produce seizure-freedom.

Anticonvulsant drugs prescribed to all children with epilepsy with onset in later childhood differed from the phenobarbital-dominated treatment regime in the neonatal period. With one exception the therapy was initiated as a monotherapy, particularly with levetiracetam (n=13), lamotrigine (n=9), oxcarbazepine (n=8), sultiame (n=6) and valproic acid (n=6) (**Figure B**). While nearly half of these patients (47.6%, 10/21) were also treated with a monotherapy (3 patients changed the drug type) at the time of the study, 11 patients (52.4%) had to be escalated to anticonvulsant drug polytherapy receiving two or more anticonvulsant drugs and therefore fulfilled the criteria of drug-refractory epilepsy. Seizure freedom for more than 6 months was only achieved in six cases (28.6%) with seizure onset after the neonatal period. This leaves 15/21 (71.4%) cases with recurrent seizures.

### Motor outcome

We further investigated the motor, cognitive and psychosocial outcome of children with PIS and seizures. In the total cohort, motor development was delayed in 47.8% (22/46), and motor delay was more frequent in those with epilepsy-onset after the neonatal period (73.7%, 14/19) than in those with acute symptomatic seizures in the newborn (29.6%, 8/27). CP was present in 52% of all cases (26/50) and in this subgroup motor delay occurred in 61.5% of cases (16/26). The prevalence of CP was higher in cases with seizure with onset in later childhood than in those with first seizures in the neonatal period (66.6% (14/21) versus 41.4% (12/29)). Still, most children with CP (84.6%, n=22/26) had no or only minor motor function impairment, with GMFCS levels I-II. Four patients had GMFCS III and, thus, moderate motor deficiency. No patient had high motor deficiency (>GMFCS III). The four patients with GMFCS III had cranial MRI findings of a unilateral MCA PIS. Almost half (46.2%) of the children with CP developed contractures and most of these required Botulinum toxin A injections (75%, 9/12) and/or surgical interventions, with tenotomy in 3 cases. Twenty patients (76.9%) required orthoses, and most (76%, 38/50) received physiotherapy. When correlating motor outcome with the time of seizure onset, there was a significant correlation between late-onset seizures and the presence of CP (p=0.036, Mann-Whitney-U-Test).

### Cognitive and psychosocial outcome

Cognitive function was assessed using standardized tests in 21 patients, while 11 patients were too young for testing, and further 17 custodians did not consent to standardized testing (**Table 1**). The test applied depended on the (developmental) age of the individual and the speech development. Developmental testing with Bayley Scales of Infant Development II (BSID-II) in 14 patients revealed a mean mental developmental index of 87.6 (range 68-103). The mental developmental index was below average in eight (57.1%) and average in six children (42.9%). In addition, the mean intelligence quotient (IQ) was 81 (range 77-86) in three children using the K-ABC, 83.2 (range 69-96) in 5 children using the WISC-IV. In summary, eight patients (38%) had an average cognitive ability, while it was below average in 61.9% (13/21) (61.9%) of the cases. Since parents of 17 cases decided not to have their children undergo formal cognitive testing, we categorized the cognition based on clinical assessment data in those with no cognitive testing data available. Taking the sum of clinical (17 patients) and formal evaluation (21 patients), 61.5% patients (24/39) had normal cognition.

Speech development was evaluated formally in 37 patients; 10 patients were too young for evaluation. Only 13.5% cases (5/37) had normal speech outcome at a mean age of 5.8 years. Most children (78.3%, 29/37) had a moderate speech delay at an average age of 8.7 years, and 8.1% of cases (3/37) showed a severe speech delay at an average age of 11.3 years and were equipped with a speech-generating device.

Special education was required in nursery school in 58.4% (23/40) of the children; 10 children did not attend kindergarten or lacked detailed information on educational support. In those 23 children old enough to attend school eight patients (34.8%) attended regular school, while 12 patients (52.2%) required special education programs due to intellectual disability. For 3 patients the school type remained unknown.

## DISCUSSION

The goal of this study was to characterize the neurodevelopmental outcome and the rate of developping recurrent seizures and refactory epilepsy in 50 children with arterial PIS, which might help to improve counselling for parents. The male predominance found in our cohort is in accordance with other studies^9,10,15.^ Extended thrombophilia screening revealed predisposition genetic factors in 52% of the patients with genetic testing, while the cause remained unclear in the remaining 25 children. Due to the small tested cohort, it remains unclear whether the prevalence of thrombophilia in children with PIS exceeds the prevalence in the normal population and thus represents a risk factor. Previous studies did not show a clear correlation^22,23^. Surely, the considerable progress in genetic and further including autoimmune testing will need to close this gap in the future to provide explanations in the counseling setting.

We found that the overall outcome of the children with PIS was influenced more by epilepsy severity and cognitive function than by motor function limitations. Motor delay or deficiency in line with CP affected half of the cohort, but motor impairment was rather mild with low GMFCS levels of I-II in most (84.6%) children. The presence of CP in 52% of all cases is in line with previous report^13,19,20^. We found motor delay to be more frequent in those with acute symptomatic seizures in the newborn than in subjects with onset in the later childhood (73.7% versus 29.6%).

When considering an extended cohort of 69 patients, i.e., also those 19 patients with PIS that lacked seizures at the time of the study, treatment for other reasons at our center and a mean age of 5.8 (range 0.4-17.4) 72.5% of this extended cohort developed seizures within the observation period of the study, which included 50 children at a mean age of 7.1 years. These 19 children without seizures were not further evaluated in the study. Of the 50 patients with PIS and seizures, 58% had first seizures in the neonatal period, while the seizure onset in the other children occurred at any age with a mean of 4 years. The prevalence of seizures with onset in later childhood in our cohort was rather high with 42% when compared to other studies reporting rates of 9-39%^7,10,13,14,24^. This may be in part due to selection bias through the exclusion of asymptomatic children with PIS. Additionally, due to the retrospective nature of the study and the database search with keyword search there might be a systmatic bias

Prospective studies are needed to provide a more accurate representation.

A total of 36% of cases with PIS and seizures had drug-refractory epilepsy. The course of epilepsy appeared to be more severe in those with later epilepsy-onset, albeit the limitation of the retrospective nature of the study with a restricted tracking of seizures throughout life. An encouraging 51.7% of cases with acute symptomatic seizures in the newborn were seizure-free in life, at the time of the study, while 24.1% had developed drug-resistant epilepsy. Of those with epilepsy with onset in later childhood, more than half required polypharmacotherapy and 52.4% were drug-resistant. These patients were further assessed for epilepsy surgery in our institution and 3 of the 21 patients (14.3%) received hemispherectomy. 26.8% of the children with epilepsy with onset in later childhood became seizure free with anticonvulsive therapy. This result is in line with the results reported for other structural epilepsies^25^ and the 58% reported for children with epileptic spasms after a perinatal stroke by Lockrow et al.^17^. The adequate treatment for epilepsy after a perinatal stroke is still unclear and further studies especially for the rate of seizure freedom are needed. Fox et al^16^. reported that acute symptomatic seizures in the newborn triple the risk of remote seizures. In accordance with Wusthoff et al^14^. we could not find a higher risk for structural epilepsy for children acute symptomatic seizures in the newborn. 48.2% of these children developed recurring seizures, albeit cases with later recurring seizures may have been missed due to the limited observation period.

The cognitive function was below average in 15/39 of children with PIS and seizures and a large number of children required special education in kindergarten and school. 61.5% patients (24/39) had normal cognition. It is discussed that there is a negative effect of seizures on cognitive performance in children with PIS,^5,16^ but we could only find an insignificant correlation in our cohort (p=0.099). Most children needed intensive physiotherapy and speech therapy; however, participation in life was usually age-appropriate.

In conclusion the main findings of our study are the high rate late-onset of epilepsy in our cohort and the low rate of seizure freedom in children with epilepsy with onset in later childhood. Acute symptomatic seizures in the newborn show a good response to AED treatment, especially phenobarbital and a better cognitive outcome than children with later onset of seizures. Therefore, an early diagnosis, follow-up examinations, adequate therapy and counseling of parents already in the neonatal period are important.

## Data Availability

The data is only available on request from the authors.

## Acknowledgements

We thank all the patients and their families. This work was supported by the German Research Foundation (DFG, SFB1315) and the Charité – Universitätsmedizin Berlin. JS is participant in the BIH-Charité Clinical Scientist Program funded by the Charité - Universitätsmedizin Berlin and the Berlin Institute of Health.

## FIGURES AND TABLES

**Figure 1.**
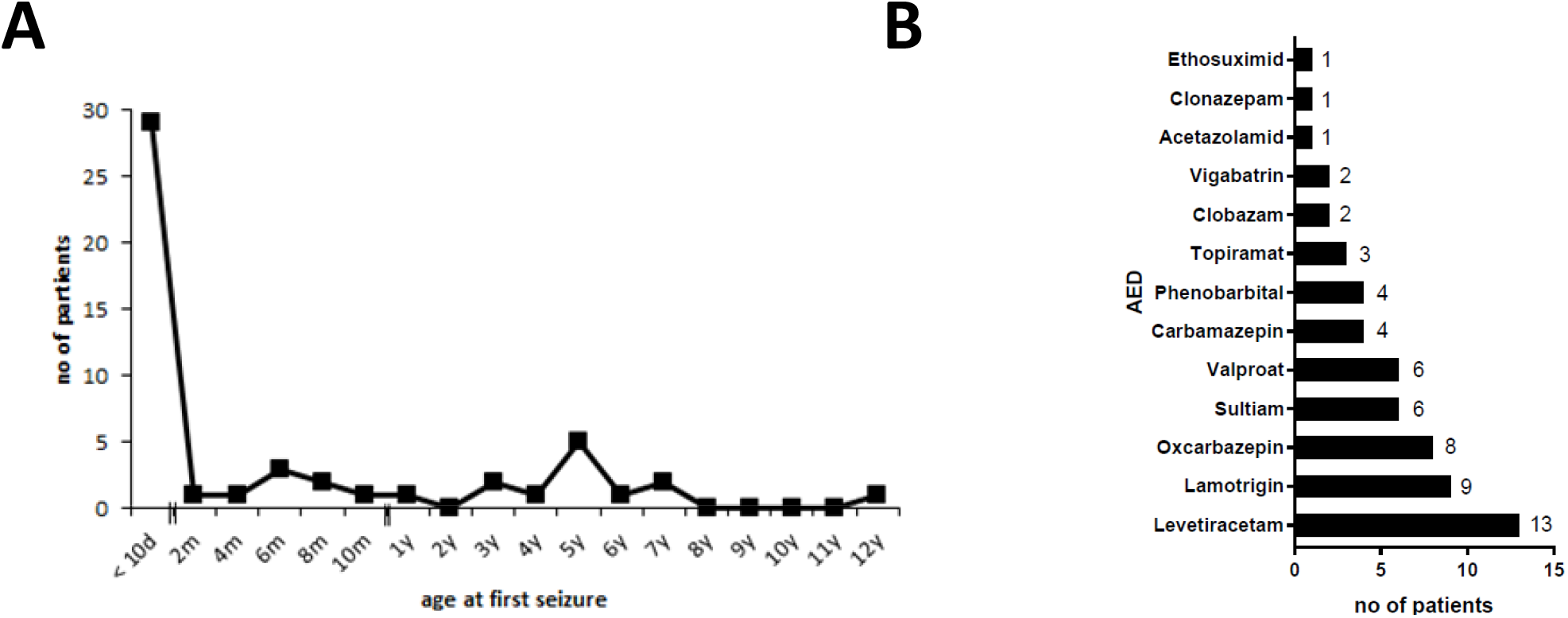
Outcome of cohort with arterial PIS and structural epilepsy. (A) Age at first seizure, (B) Number and type of drugs used as AED for non-neonatal onset

**Table 1. Clinical, radiological and laboratory variables of cohort.**

## REFERENCES

1. Lynch, J. K. & Nelson, K. B. Epidemiology of perinatal stroke. Curr Opin Pediatr 13, 499–505 (2001).

2. Schulzke, S., Weber, P., Luetschg, J. & Fahnenstich, H. Incidence and diagnosis of unilateral arterial cerebral infarction in newborn infants. J Perinat Med 33, 170–175 (2005).

3. Rattani, A. et al. Incidence of Epilepsy and Associated Risk Factors in Perinatal Ischemic Stroke Survivors. Pediatric Neurology 90, 44–55 (2019).

4. De Haan, T. R., Langeslag, J., van der Lee, J. H. & van Kaam, A. H. A systematic review comparing neurodevelopmental outcome in term infants with hypoxic and vascular brain injury with and without seizures. BMC Pediatr 18, 147 (2018).

5. Raju, T. N. K., Nelson, K. B., Ferriero, D. & Lynch, J. K. Ischemic Perinatal Stroke: Summary of a Workshop Sponsored by the National Institute of Child Health and Human Development and the National Institute of Neurological Disorders and Stroke. Pediatrics 120, 609–616 (2007).

6. Sorg, A.-L. et al. Incidence Estimates of Perinatal Arterial Ischemic Stroke in Preterm- and Term-Born Infants: A National Capture-Recapture Calculation Corrected Surveillance Study. Neonatology 1–7 (2021) doi:10.1159/000514922.

7. Fitzgerald, K. C., Williams, L. S., Garg, B. P. & Golomb, M. R. Epilepsy in Children With Delayed Presentation of Perinatal Stroke. J Child Neurol 22, 1274– 1280 (2007).

8. Ballantyne, A. O., Spilkin, A. M., Hesselink, J. & Trauner, D. A. Plasticity in the developing brain: intellectual, language and academic functions in children with ischaemic perinatal stroke. Brain 131, 2975–2985 (2008).

9. Kirton, A. et al. Symptomatic Neonatal Arterial Ischemic Stroke: The International Pediatric Stroke Study. PEDIATRICS 128, e1402–e1410 (2011).

10. Grunt, S. et al. Incidence and Outcomes of Symptomatic Neonatal Arterial Ischemic Stroke. Pediatrics 135, e1220–e1228 (2015).

11. Williams, T. S. et al. Secondary attention-deficit/hyperactivity disorder following perinatal and childhood stroke: impact on cognitive and academic outcomes. Child Neuropsychology 24, 763–783 (2018).

12. Srivastava, R., Shaw, O. E. F., Armstrong, E., Morneau-Jacob, F.-D. & Yager, J. Y. Patterns of Brain Injury in Perinatal Arterial Ischemic Stroke and the Development of Infantile Spasms. J Child Neurol 088307382098605 (2021) doi:10.1177/0883073820986056.

13. Lee, J. et al. Predictors of outcome in perinatal arterial stroke: A population-based study. Ann Neurol. 58, 303–308 (2005).

14. Wusthoff, C. J. et al. Risk of Later Seizure After Perinatal Arterial Ischemic Stroke: A Prospective Cohort Study. PEDIATRICS 127, e1550–e1557 (2011).

15. Chabrier, S. et al. Multimodal Outcome at 7 Years of Age after Neonatal Arterial Ischemic Stroke. J Pediatr 172, 156-161.e3 (2016).

16. Fox, C. K., Glass, H. C., Sidney, S., Smith, S. E. & Fullerton, H. J. Neonatal seizures triple the risk of a remote seizure after perinatal ischemic stroke. Neurology 86, 2179–2186 (2016).

17. Lockrow, J. P., Wright, J. N., Saneto, R. P. & Amlie-Lefond, C. Epileptic Spasms Predict Poor Epilepsy Outcomes After Perinatal Stroke. J Child Neurol 34, 830–836 (2019).

18. Fluss, J., Dinomais, M. & Chabrier, S. Perinatal stroke syndromes: Similarities and diversities in aetiology, outcome and management. European Journal of Paediatric Neurology 23, 368–383 (2019).

19. Clive, B. et al. Epidemiology of neonatal stroke: A population-based study. Paediatrics & Child Health 25, 20–25 (2020).

20. Biswas, A., Mankad, K., Shroff, M., Hanagandi, P. & Krishnan, P. Neuroimaging Perspectives of Perinatal Arterial Ischemic Stroke. Pediatric Neurology 113, 56–65 (2020).

21. Palisano, R. et al. Development and reliability of a system to classify gross motor function in children with cerebral palsy. Developmental Medicine & Child Neurology 39, 214–223 (2008).

22. Curtis, C. et al. Thrombophilia risk is not increased in children after perinatal stroke. Blood 129, 2793–2800 (2017).

23. Lehman, L. L. et al. Workup for Perinatal Stroke Does Not Predict Recurrence. Stroke 48, 2078–2083 (2017).

24. Suppiej, A. et al. Pediatric epilepsy following neonatal seizures symptomatic of stroke. Brain and Development 38, 27–31 (2016).

25. Geerts, A. et al. Course and outcome of childhood epilepsy: A 15-year follow-up of the Dutch Study of Epilepsy in Childhood: Course and Outcome of Childhood Epilepsy. Epilepsia 51, 1189–1197 (2010).

26. Bosenbark, D. D. et al. Clinical Predictors of Attention and Executive Functioning Outcomes in Children After Perinatal Arterial Ischemic Stroke. Pediatric Neurology 69, 79–86 (2017).

